# A realistic way to investigate the design, and mechanical properties of flow diverter stents

**DOI:** 10.1101/2021.03.23.21253935

**Authors:** P. Velvaluri, M. S. Pravdivtseva, J. Hensler, F. Wodarg, O. Jansen, E. Quandt, J. B. Hövener

## Abstract

**Purpose:** Braided flow diverters (FD) are highly sophisticated, delicate, and intricate mechanical devices used to treat intracranial aneurysms and thus saving lives. Testing such devices in vitro, however, remains an unsolved challenge. Here, we evaluate methods that access flow, design, and mechanical properties in vitro.

**Methods:** Flow properties, cell porosity, and cell area were evaluated by placing FDs in patient-derived, 3D printed models of human vasculature. 4D flow MRI was used to measure fluid dynamics. Laser microscopy was used to measure porosity and cell area with the top of aneurysm sac cut off for the model. New testing methods were developed to investigate the bending, circumferential, and longitudinal radial force continuously over varying diameters.

**Results:** The placement and flow properties of the FD in the vasculature models were successfully measured by MRI, although artifacts occurred. The setup to measure porosity and cell area inside of the model proved successful. The newly discussed methods allowed us to measure the indicated forces, to our knowledge for the first time, continuously.

**Conclusion:** Modern and specifically tailored techniques, some of which were presented here for the first time, allow detailed insights into the flow and mechanical properties of braided flow diverter stents.

## 1. Introduction

An intracranial aneurysm is an abnormal dilatation of the brain arteries with a 30-day mortality rate of 45 % in the case of hemorrhagic stroke [1]. Therefore, it is vital to identify and treat aneurysms that may rupture. Different approaches can be used to treat aneurysms [2]. One of the new and successful aneurysm treatment methods is reconstructing a vessel using stand-alone braided flow diverter (FD) devices [3–5]. FDs were found useful for the treatment of aneurysms with complex anatomy [6]. Despite numerous and often successful FDs applications, complications during and after FDs placement still occur, resulting in, e.g., a (delayed) aneurysm rupture [7,8]. An aneurysm occlusion after the deployment of FD occurs via two main biological mechanisms: the formation of intra-aneurysmal thrombosis caused by flow disruption or the growth of endothelial cells on the device struts itself [9]. The understanding of both mechanisms is crucial to reduce the probability of failed aneurysm treatment using FDs. Various device properties (design and mechanical) may influence the biological mechanisms, thereby changing the clinical outcomes.

Previously, mechanical properties such as circumferential and longitudinal radial force [10,11] of intracranial stents were discussed. However, these properties were evaluated only at particular stent diameters. The circumferential radial force was calculated at 50 % [10] and 85 % [11] of the stent diameter. The longitudinal radial force was reported in both studies at half of the stent diameter. It is known that the radial forces vary with diameter, as the force that the stent exerts depends on the amount of compression. The human anatomy, on the other hand, does not provide vessels with a single diameter, e.g., the nominal diameter anterior communicating artery (ACoA), where 35 % of saccular aneurysms occur [12], varies from 0.9-2.4 mm for an adult human [13]. In the brain, the blood vessels gradually decrease their diameter from the circle of Willis until they reach capillaries, which are only a few microns in diameter. Therefore, varying diameter over an artery length of 20-30 mm (standard stent lengths) is very common. So, there is a clear need to understand how these devices exert forces over a range of diameters.

Moreover, porosity and cell-area of the braided FDs are essential design properties. The porosity in braided FDs depends on multiple parameters such as compression length and bend angle, among others [14]. The porosity itself is an essential characteristic of the stent design because it can influence the flow into the aneurysm [15]. These properties were investigated in the past, but only on simple, straight tubes [10,16] for braided FDs. When placed in vivo, the stents are bent, affecting the porosity and cell-area dramatically. Therefore, there is a need to evaluate these design parameters when placed in complicated aneurysm geometries.

Meanwhile, there are several braided FDs that are available in the market [17,18]. It is difficult for a neuro-interventionalist to compare the FDs objectively as in one clinical case, one can only implant one FD, and every clinical case is (anatomically) different; one cannot really compare the effect of one FD to another. For this purpose, we propose a practical approach to evaluate FDs for their design and mechanical properties. We present methods to measure the circumferential and longitudinal radial force, bending force as a continuous function of the diameter. Furthermore, we measure the porosity and cell-size not in straight tubes, but realistic configuration with 3D printed, patient-derived models of human vasculature. We used 4D flow MRI to access the flow dynamics with and without stents and verify the stent placements. As some of the testing methods are not standardized, they are described in detail here for reproducibility. We tested this battery on three commercial FD stents (P64 flow modulation device, Phenox; Derivo embolization device, Acandis; Silk, Balt extrusion) to see if the proposed tests work and not to compare the FDs.

## 2. Methods

### 2.1 Endovascular stent placement and flow evaluation

#### Aneurysm model

A patient-derived aneurysm model of the left internal carotid artery was produced based on 3D rotational angiographic data (3D RA, Figure 1a) acquired from a patient (female, between 50 - 55 years old). It was obtained as a part of a clinical routine using an X-ray angiographic system (Allura Xper FD20/10 biplane, Version R7.2.9, Philips Healthcare, Netherlands). The data were anonymized before further processing. The protocol to produce the model comprised: 1) segmentation of a vascular lumen from the 3D RA dataset (Figure 1a); 2) optimization of the resulting lumen (removing artifacts: closing holes, separating vessels, (Figure 1b) and 3) model preparation for printing (adding external wall and connectors for integration into the flow loop; 4) printing a digital model with stereolithographic 3D Printer (Clear photoreactive resin, Form 2 Formlabs, USA). A detailed protocol of the model fabrication is submitted for publication. The study was approved by the Ethics Committee at the Faculty of Medicine, Kiel University. Four aneurysm models were printed to place the three FDs and one control. The FDs were placed into the models (Figure 1c) by experienced neuroradiologists.

**Figure 1:**
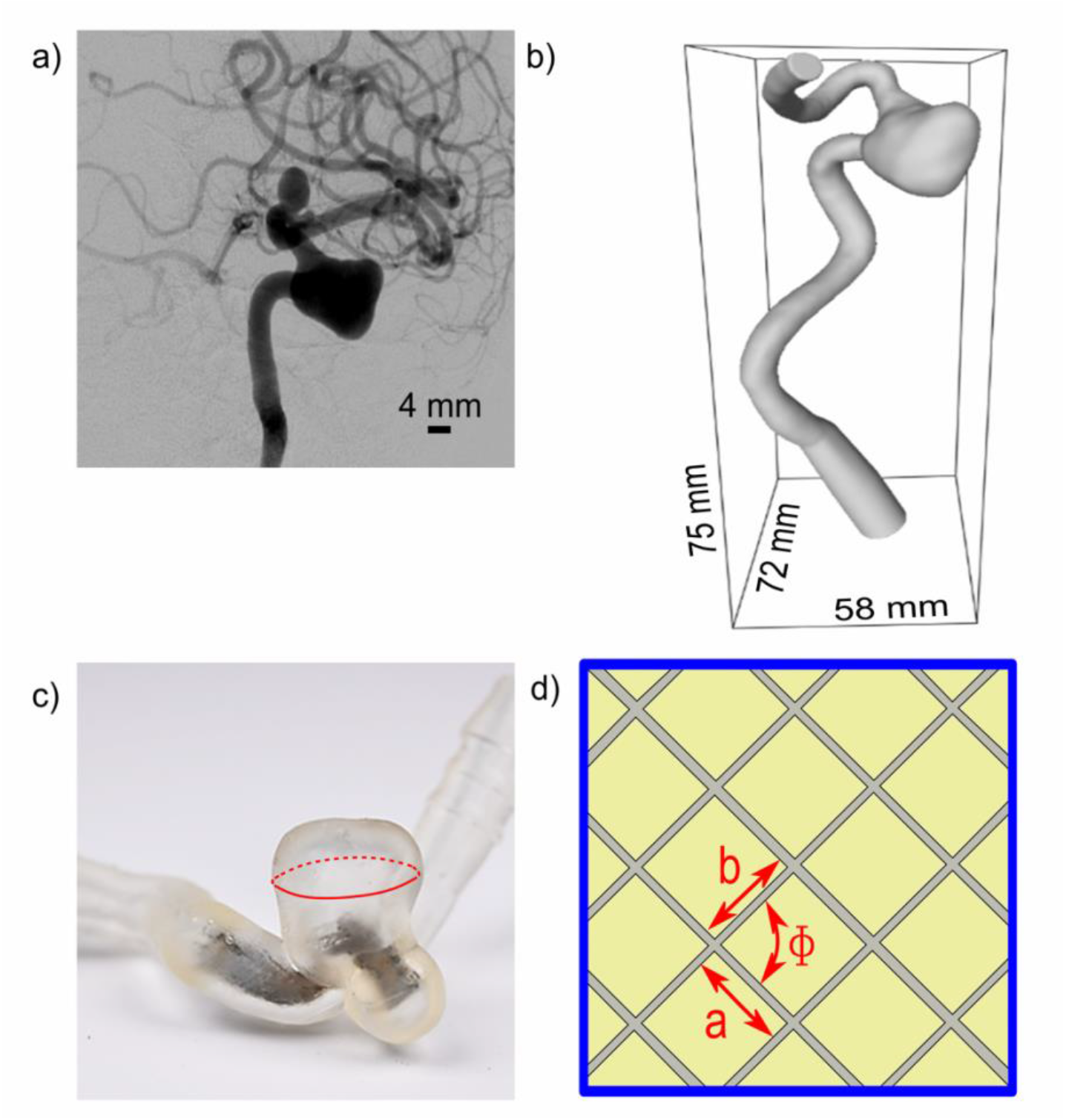
(a) 2D digital subtraction angiography data (DSA) of a patient (female, between 50 - 55 years old) with left internal carotid artery aneurysm. (b) A vascular lumen was segmented from 3D RA, and the side branches were removed from the original data to simplify the model. (c) Photo of the 3D printed patient-derived model with example deployed stent. The aneurysm sac was cut (red line); this way, the stent surface was visible, and it was subjected to the laser microscope imaging. (d) Illustrative laser microscope image of the stent surface for the porosity and the cell-area calculations. The porosity was calculated as the ratio to open cell-area (sum of yellow areas) to total area (blue), and the cell area of the parallelograms was calculated using the parameters (a, b, ϕ) obtained from the image.

#### Flow imaging

The flow in aneurysm and vessel was detected by flow imaging using 4D flow MRI. The models were integrated into a flow-loop (40:60 glycerol-water mixture, a flow rate of 4.4 ml/s Simuflow 1000, Schelley Systems, USA). Flow-sensitive magnetic resonance imaging (MRI) (4D flow MRI, [19]) was performed on a 3 T whole-body MRI system (Achieva, Version R5.3, Philips Healthcare, The Netherlands, 32-channel head coil). 4D Flow MRI was performed using 3D T1 weighted, turbo field spoiled gradient echo (field of view: 40 x 40 x 30 mm^3^, acquired voxel size: 0.63 x 0.63 x 0.63 mm^3^, echo time: 5.1 ms, repetition time: 12 ms, flip angle: 8°, velocity encoding: 110 cm/s, temporal resolution 40 ms). The resulting velocity distribution was analyzed in the aneurysm sac and parental vessel. The regions of interest were created manually based on the vascular surface, segmented from the intensity-weighted maximum-velocity data obtained by 4D flow.

### 2.2 Porosity and cell-area evaluation

The upper part of the aneurysm sac of the patient-derived models was manually removed (redline, Figure 1c). Thus, making the deployed stent’s surface visible and was subjected to the laser microscope (Model VK-X260K, Keyence, Germany) imaging. The illustrative image shows the deployed 3D stent surface as a 2D projection (Figure 1d). The length of the two adjacent facets (a, b) and angle (*ϕ*) between them were measured manually (using ImageJ, open-source, Figure 1d) and was calculated using equation 1. The area was calculated for different cells in a single image, and the mean and standard deviation were evaluated. Porosity was assessed as a ratio of open areas (Figure 1d, yellow) to the image’s total area (Figure 1d, blue square), as given in equation 2. After the analysis, the FDs were gently removed from the patient-derived models for further mechanical testing.

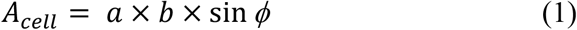

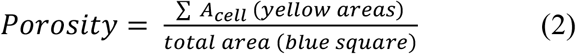

### 2.3 Mechanical testing

#### 2.3.1 Circumferential radial force (RF_c_)

RF_C_ was measured with an in-house-built setup (Figure 2a) during the expansion of the stent. The FDs were 1) crimped to a diameter of 2.2 mm (approx. half of the free diameter), 2) deployed into the apparatus (red dotted circle), and 3) gradually released till free diameter (4– 4.3 mm). The crimping device was made in-house after slightly modifying an available patent by Motsenbocker et al. (US007069794B2) [20]. The device consists of 12 radial blades that are pivoted to move as freely as possible. The ends of the radial blades are connected to a force-ring, which in turn is connected to an assembly of force sensor (100 N, Disynet GmbH, Germany) and linear actuators (M-229.26S & L-306.011100, PI miCos GmbH, Germany). So, when a stent expands, it exerts force on the radial blades-force ring-sensor. Thus, the RF_C_ was measured as a function of the stent diameter during expansion. Details about the correction factor to accommodate the friction is discussed in the supplementary (S. Figure 1).

**Figure 2.**
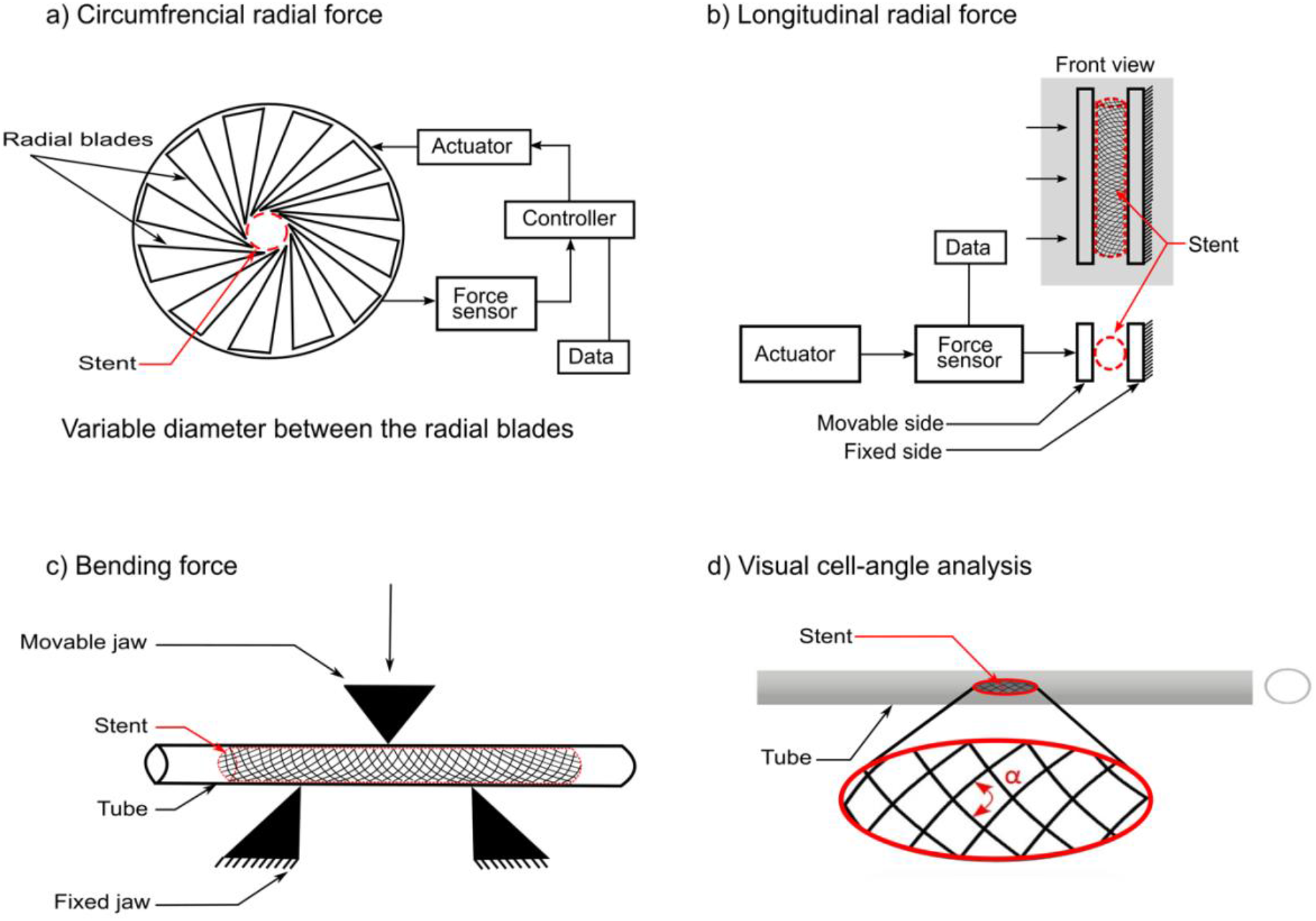
(a) Scheme of the in-house-built setup to measure the circumferential radial force (RFC). An FD (red dotted circle, red arrow) was crimped to a diameter of 2.2 mm and inserted between radial blades of the apparatus. The FD was gradually expanded in the apparatus. The force exerted on radial blades was recorded as a function of stent diameter. (b) Scheme of the setup to measure the longitudinal radial force (RFL). The front view (a grey area) and a top view of the apparatus are shown. The stent (red dotted line) was placed between two parallel plates; one was fixed and another movable. The stent was compressed longitudinally with a movable plate to a displacement of 2.2 mm from its initial diameter. The force was recorded as a function of the plate displacement. (c) Scheme of the setup to measure the bending force (BF). The stents were deployed in the 3D printed tubes of various diameters (3–4.25 mm; with intervals of 0.25 mm). The tubes were bent to a 14 mm radius of curvature, and the force required to bend was measured. (d) The light microscope was used to generate images of the stent surface for visual cell-angle deployed in 3D printed tubes. Each tube had a hole (red outline) to expose the stent surface, and the angle of the parallelogram (α) of the stent surface was measured.

#### 2.3.2 Longitudinal radial force (RF_L_)

RF_L_ was measured with an in-house-built setup (Figure 2b) during stent compression. The device’s setup is shown in the line diagram, the linear actuator (M-229.26S, PI miCos GmbH) coupled with a force sensor (100 N, Disynet GmbH) was connected to the movable plate. The front view (yellow) and a top view of the apparatus can be seen. The FD (red dotted line) was placed between two parallel plates (fixed and movable). The FDs were compressed longitudinally to a noncircular cross-section from 2 mm (approximately half of the diameter) from the free diameter (4–4.3 mm). Thus, the RF_L_ was measured as a function of the movable side displacement.

#### 2.3.3 Bending force (BF)

BF was evaluated using a standard 3-point bending method (Figure 2c). FDs were deployed in 3D printed flexible tubes of various diameters (3-4.25 mm, with increments of 0.25 mm, 3-D printed with Flexible Photoreactive Resin, Form 2, Formlabs, USA). The tubes with FDs were bent to a 14 mm radius of curvature with a setup of modified holders (movable and fixed jaw) in tensile tester (100 N, ZwickRoell Z0.5, Germany) (Figure 2c). The required BF to bend each tube was detected. The normalization of the BF was done for FD1, FD2, with the corresponding value of FD3 at each diameter. Additionally, a different tube (3-4.25 mm, with increments of 0.25 mm) was designed with a small window at the center (8 mm) (Figure 2d, red oval). Such opening provided optical access to evaluate the cell-angle between stent struts (α). The cell-angle measurement was performed using the light microscope (Eclipse L200D, Nikon, Japan). As each image had many cells, the visual cell-angle measurements were reported with a mean and standard deviation.

All measurements (RF_C_, RF_L_, and BF) were performed at body temperature (37°C). All the measurements were performed three times for each device per testing method, and the corresponding standard deviation was shown with error bars.

### 2.4 Endovascular stents

Three flow diverter stents (FD1-3, details below) with a free diameter of 4.0–4.3 mm and length of approximately 30 mm were considered as trail stents. All dimensions of the devices were chosen by experienced neuroradiologists for the considered aneurysm geometry.

FD1: P64 flow modulation device, Phenox, Germany, 64-NiTi wire braid, visibility through two helical strands along with the entire length, and eight proximal markers [21].

FD2: Derivo embolization device, Acandis, Germany, radiopaque NiTi composite wires with platinum core, and three Platinum-Iridium X-ray markers on both ends [22].

FD3: Silk, Balt extrusion, France, 48 NiTi wires, four radio-opaque markers over the entire length, eight extra Platinum smaller wires in the stent [23].

## 3. Results

### 3.1 Endovascular stent placement and flow evaluation

Flow-sensitive MRI of the 3D printed models of human vasculature with and without stents was successfully acquired (Figure 3). Signal voids likely due to metal artifacts were observed predominantly close to the stents’ surface, such that the MRI signal appeared to be strongly reduced in these areas (Figure 3a, yellow arrows). A qualitatively substantial flow jet entering the aneurysm sac was observed without implants, and in the presence of FDs, the flow jet was eliminated (Figure 3b).

**Figure 3.**
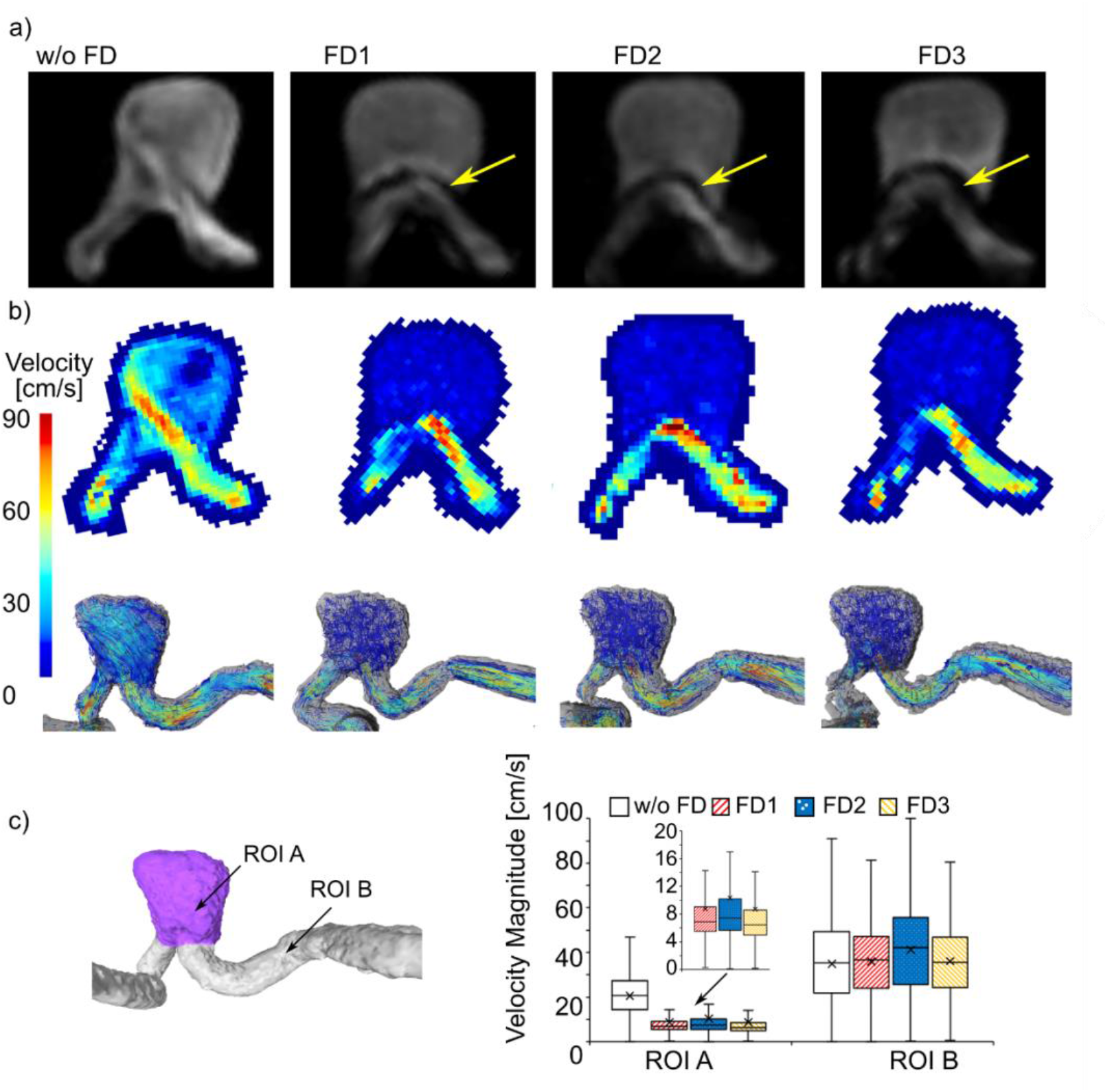
(a) Magnitude, (b) velocity, and (c) pathline reconstructions of the flow through the printed model with and without FDs, acquired with 4D flow MRI (protocol 1). (a) Note the signal voids, yellow arrows, indicating the positions of the stents. (b) The velocity maps of the same slice indicate a significant diversion of the flow. (c) As expected, a quantitative analysis of the data found that the mean velocities were dramatically reduced in the aneurysm sac (ROI A), while it was constant in the parent vessel (ROI B).

The mean velocity was analyzed in the aneurysmal sac and parent vessel (ROI A-B Figure 3c). In the aneurysmal sac, the mean velocity was reduced for all stents after their implantation (s. Table1, without FD: 20.53±13.53 cm/s versus 8.76±8.45 cm/s, 10.35±11.32 cm/s and 8.71±9.02 cm/s for FD1-3 respectively). In the parental vessel, it was almost constant (s. Table1, without FD: 34.89±20.12 cm/s, with FD 1-3: 36.03±17.54 cm/s, 41.27±21.33 cm/s, 36.12±16.74 cm/s, respectively). Note that a large standard deviation is expected, mostly for two reasons: 1. developed flow has a parabolic profile in a circular cross-section with a maximum value at the center and a zero value at the wall; 2. The vessel diameter is not constant across the model, resulting in a change in the velocity to maintain the total flow. Overall, the data indicate a sufficient flow diversion for all stents, suggesting that the stent placement was successful.

### 3.2 Porosity and cell-area evaluation

The upper part of the aneurysm model was cut-off (yellow arrow) to examine the stent surface (green rectangle, Figure 4a). The area between stent wires (cell-area) and porosity was evaluated at proximal (Pos. 1), central (Pos. 2), and distal (Pos. 3) positions of the stent (blue arrows). The laser microscope image was analyzed to obtain the cell parameters (a, b, and *ϕ*) (Figure 4b). The stent porosity (Figure 4c) of the FD surface was calculated using equation (2) at three different positions (Pos. 1-3) from these pictures. The porosity of FD1-2 decreases from Pos. 1 to Pos. 3 by approximately 5.5 %, whereas for FD3, it increases by 4 %. The cell-area of the FD surface were calculated using equation (1) at the same positions (Pos. 1-3) (Figure 4d). A slight decrease in the cell-area was observed for FD1-3. For better quantitative comparison, the cell-area at the Pos. 2-3 was normalized by the area at Pos. 1 (Figure 4e). The highest decrease in the area from Pos. 1-3 was yielded by FD1 (24 %), followed by FD2 (13 %) and then FD3 (5 %). The error bars indicate the standard deviation of the different cell-areas calculated in the same image.

**Figure 4.**
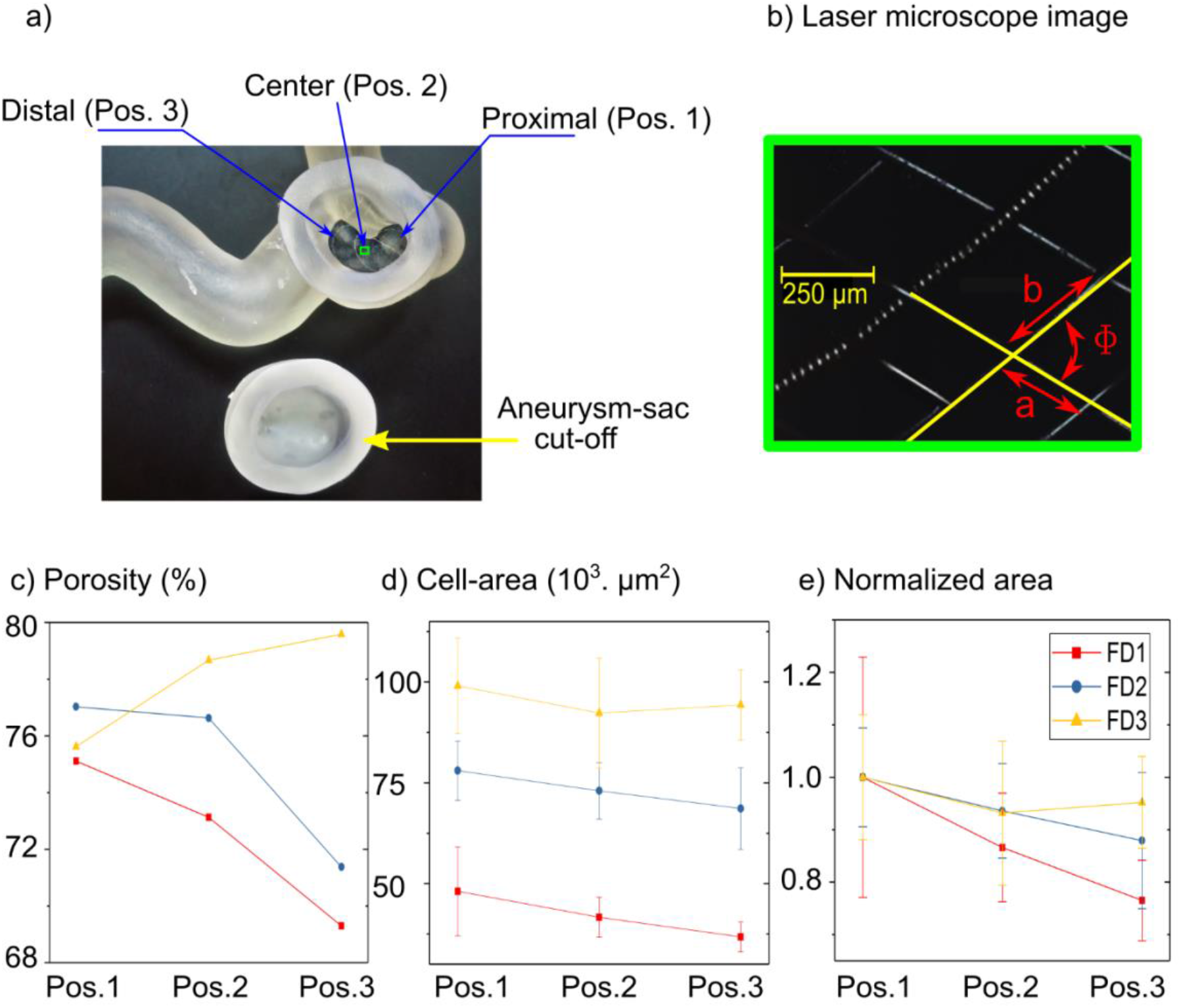
(a) Photo of the 3D printed patient-derived model with a deployed stent. The aneurysm sac was cut off (yellow arrow) to provide access for a laser microscope. The images were taken at three different positions (Pos. 1-3), as indicated by blue arrows. (b) The laser microscope image of the stent surface was used to calculate porosity and the cell-area using the parameters (a, b, Ø). (c) Resulting porosity at positions 1 -3. Interestingly, the porosity was reduced for FD1-2 from Pos. 1 to Pos. 3. In contrast, for FD3, the porosity increased for the same. (d) Measured cell-area at three different positions (Pos. 1 -3). Note that the cell area was reduced slightly from proximal to distal positions. (e) The calculated cell-area normalized to value at Pos. 1, however, showed that the decrease from Pos.1 to Pos.3. The error bars indicate the standard deviation for cell-areas calculated on the image obtained at a specific position.

### 3.3 Mechanical testing

#### Circumferential radial force (RF_C_), Longitudinal radial force (RF_L_), and Bending force (BF) analysis

The RF_C_ was measured as FDs expanded to their free diameters, which was slightly different for each FD (4.0–4.3 mm). The RF_C_ exerted by all the FDs during expansion was very similar (Figure 5a). Moreover, the FDs showed two varying regimes of expansion, which are marked in yellow. The lower diameter regime has a negative slope of 2.38 N/mm, an order of magnitude higher than the slope at a higher diameter regime (0.20 N/mm).

**Figure 5.**
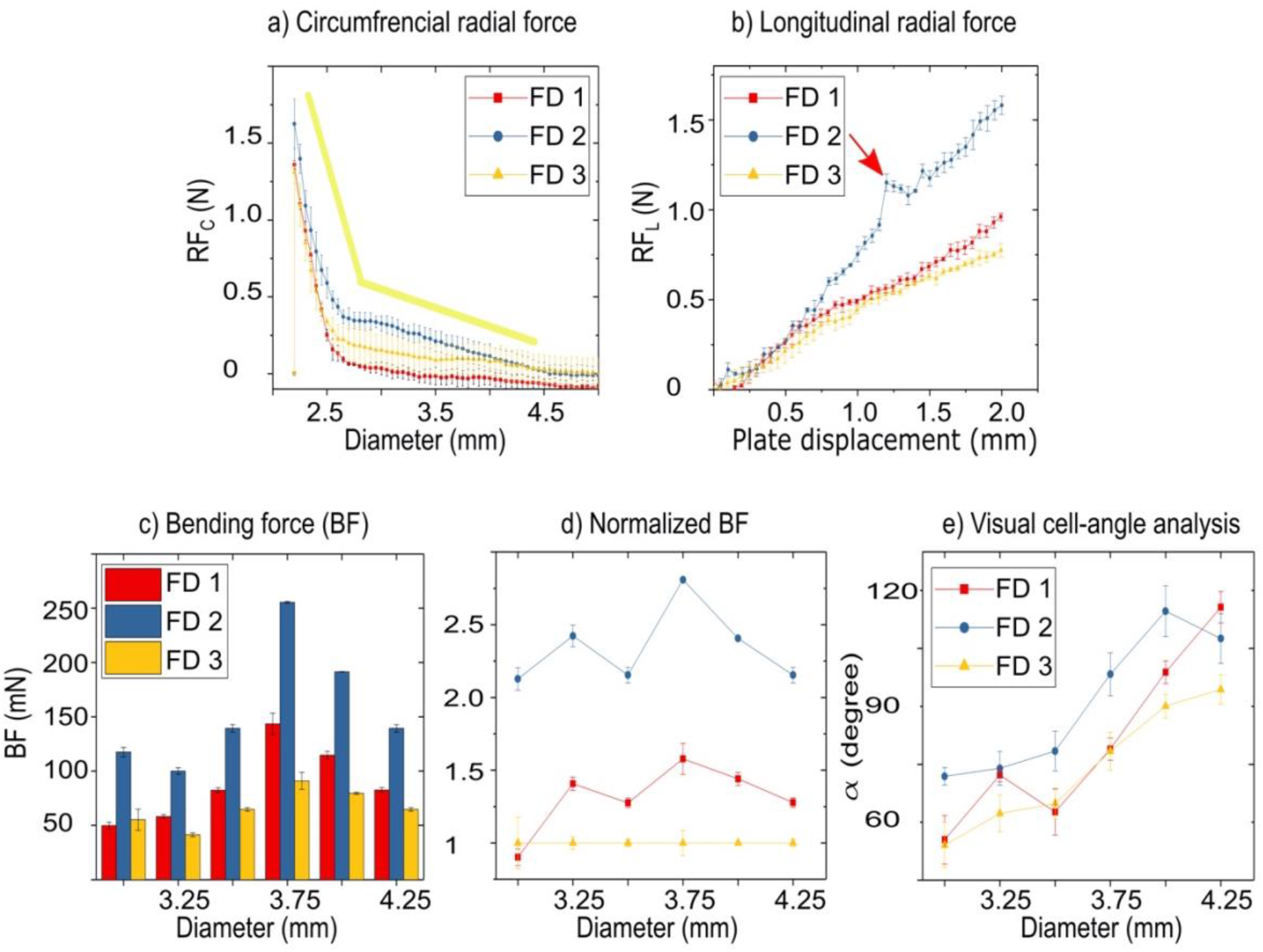
(a) Circumferential radial force (RFC) exhibited two different linear regimes during the expansion (indicated in yellow), with FD2 exerting the most and FD3 exerting the least force. (b) Longitudinal radial force (RFL) was increasing approximately linearly during the compression. A rapid increase feature (red arrow) is consistent with all the three repetitions, suggesting that it might be caused by the particular design of the FD2 stent. (c) Bending force (BF) required to bend FD1 – FD3 to a bending radius of 14 mm when deployed in tubes with diameters 3 – 4.25 mm (three-point-bending test). (d) Normalized BF for FD1 and FD2 with the value of FD3 at every diameter is shown. Note that the maximum BF required for all the FDs is 3.75 mm diameter and gradually decreases for the lower and higher diameters. From the normalized graph, it is evident that the force required to bend FD2 was more than twice compared to FD3, and for FD1, it was approximately 1.5 times higherthan FD3. Each data point was measured three times, and the error bars indicate the standard deviation. (e) Visual cell-angle analysis Figure 2) of the unit cells on the FD surface deployed in different diameters. Each diameter was imaged 3 three times, many cells were measured from each image, and the standard deviation is as indicated.

The RF_L_ was plotted as a function of plate displacement (Figure 5b). The force exerted by FD2 (1.58 N) is higher by a factor of two compared to FD1 (0.96 N) and FD3 (0.77 N) at a plate displacement of 2 mm. A rapid increase of the RF_L_ at 1.2 mm of FD2 compression (red arrow) was reproducible over three measurements, suggesting that this could be arising due to the particular design of FD2.

The BF was measured as a force required to bend the flexible tubes with deployed FD1-3 (Figure 5c) to a radius of 14 mm. Note that all the FDs showed a maximum BF at a 3.75 mm diameter and gradually decreasing for the lower and higher diameters. The normalized BF of FD1 and FD2 with the value of FD3 at every diameter (Figure 5d) was evaluated. The force required to bend FD2 was more than double compared to FD3, and for FD1, it was approximately 1.5 times higher than FD3. Additionally, the cell-angle (α) was evaluated between the struts (Figure 3f) for FD 1-3, depending on the tube diameter (3–4.25 mm, Figure 5e). We noticed a general trend that the angle (α) increases as the diameter increases for all the FDs.

## 4. Discussion

The current high selection of braided FD devices for the treatment of aneurysms does not allow the interventionalist to pick the best device suited for the given anatomy of the patient. Our work shows a practical approach to characterize FDs and help interventionalists compare the FDs objectively.

### 4.1 Endovascular stent placement and flow evaluation

4D flow MRI allowed to access the flow diversion in the patient-derived models (Figure 3a-c). A 50-58 % reduction in mean intra-aneurysmal velocity was detected, which shows that the devices are successfully placed along the aneurysm’s neck.

However, 4D flow MRI is prone to metal-induced artifacts. Earlier, the influence of metal artifacts in the presence of the braided stents (FD1 and FD3) on flow estimation was investigated [24]. The stents were placed in the straight tubes; first, it was shown that the static background was not affected by the stents, and, second, the mean velocity magnitude was almost the same along the tube in case of parallel orientation of the stent to the magnetic field. In our study, the model was oriented along the direction of the magnetic field; thus, the stent was mostly aligned parallel to the magnetic field. Another technique, e.g., single-point 4D flow MRI [25], can be more robust toward the artifacts, but the scan times are too long for in vivo applications.

Other shortcomings of 4D flow MRI often include insufficient spatial resolution for small vessels and aneurysms, limited sensitivity to the low velocities, and non-repetitive flow patterns. Higher fields [26] and acceleration techniques [27] can overcome some of these issues; real-time MRI may be used to image non-repetitive flow [28]. The flow diversion can be investigated with higher accuracy by blood simulation or particle-image velocimetry. However, the latter works exclusively *in vitro* and has no equivalent *in vivo*. In contrast, 4D flow MRI examination can be the same for both *in vitro* and *in vivo*.

### 4.2 Porosity and cell-area evaluation

The porosity (Figure *4*a) was almost constant (70-80 %) over Pos. 1-3 for all the FDs. The variation can be due to the change in the vessel’s diameter from Pos. 1 (3.67 mm) to Pos. 3 (4.25 mm), diameters obtained from 3D model. The stent cell-area decreases from Pos.1-3 such a variation can be caused due to two reasons—first, the variation in the model vessel diameter. Second, the compression/foreshortening of the stent during placement, which can be done by the physician to achieve a denser mesh near the neck of the aneurysm.

The proposed method allowed to evaluate the porosity and cell-area in complex models. To our knowledge, they were evaluated previously using simplified models of vessels (straight tubes [10,16]). It is essential to use a laser microscope instead of a light microscope as the latter can only provide good focus only to a specific focal point. When placed in curved geometries, the stent surface can have structures at multiple focal points, making it challenging to use a light microscope. On the other hand, the laser microscope scans for the surface from an absolute minimum to maximum interval in z-direction and generates a 2D stent surface image for analysis.

This technique’s main shortcoming is that the laser scan images differ slightly compared to the actual data. The error arises as the placed stent is in 3D, and the acquired laser image is in 2D; this 2D projection varies slightly with the degree of the stent curvature. Such error scale linearly with the curvature of the stent surface. The higher the degree of bending, the larger is the error. The error was minimized in the current study by taking the scans from a small area, limiting the degree of bending. More analysis and studies are to be done to better account for these errors. The histological analysis of animal models is another way to evaluate porosity in complicated geometries [29]. Though these models give additional information about the cell-growth on the stents, their main limitation is that one animal model is required to test the placement of one stent by one interventionalist.

### 4.3 Mechanical testing

The previous studies [10,11] reported on braided FDs for RF_C_ measurements were done at a particular diameter (85 % of free diameter). In the current analysis, the RF_C_ was measured first, as a function of diameter (2.2 mm to free diameter) and second, at 85 % of free diameter (*Table 1*). The evaluation as a function of diameter could give a better estimate of the force if the FDs were deployed in a changing diameter scenario (distal to proximal), often the case in the human brain [13]. In comparison with literature (Table 1), the resulting RF_C_ is slightly less than it was reported in the previous studies with braided FDs; this could be attributed to the differences in the stent properties (e.g., geometrical, design, and material). The reliability and reproducibility of this method are trustworthy, as the error bars indicate minimal spread. Commercially RFc measuring devices (also called a crimping machine) are relatively well established. Providing the data of different flow diverters for a range of diameters instead of at a particular value can help compare different devices.

**Table 1.** Comparison of the RF_C_ and RF_L_ from previous studies on braided FDs only. As the studies were reported with stents of various dimensions, a normalized value (per length) was calculated for comparison. The current data was obtained when the stents were deformed to 85% of diameter in RF_C_ and 50 % of diameter in RF_L_, both normalized for a length of 30 mm.

| Study | Device | $RF_C$ (N/mm) | | $RF_L$ (N/mm) | |
| --- | --- | --- | --- | --- | --- |
| Krischek et al. [11] | Leo | 0.025 |  | 0.0037 |  |
| Cho et al. [10] | Leo+ | 0.065 | 0.05 | 0.013 | 0.007 |
|  | LVIS | 0.013 | 0.032 | 0.007 | 0.009 |
| Current study<sup>1</sup> | P64 (FD1) | 0.002 |  | 0.032 |  |
|  | Derivo (FD2) | 0.010 |  | 0.052 |  |
|  | SILK (FD3) | 0.007 |  | 0.026 |  |

Leo, Leo+: Balt, Montmorency, France; LVIS: Microvention, Tustin, CA, USA.
<sup>1</sup> The $RF_C$ was measured at 85 % of the diameter, i.e., 3.4 mm for FD1 and FD2; 3.85 mm for FD3. The $RF_L$ was measured at ~50 % of the diameter, i.e., 2 mm from the free diameter for all FDs.

In general, brain vessels have a noncircular cross-section; thus, it is essential to evaluate not only RF_C_ but also RF_L_. The effect of RF_L_ investigations for stent ovalization shows a similar trend between different stents. The analysis shows that the RF_L_ varies continuously with the plate compression. Compared to literature (Table 1) of braided FDs, our data has slightly higher values. The same argument can be considered as before, as the previous studies were focused on different devices. The reliability and reproducibility of these tests are good with a mean error of 0.032 N (calculated as a mean of all error bars) and with a maximum error of 0.081 N.

Moreover, we observed that the forces reduce and increase for diameters higher and lower than 3.75 mm for BF analysis. The normalized BF graph shows the force exerted between the different stents FD 1-3. The visual cell-angle at 3.75 mm diameter shows that high force is observed when α is approximately 90 degrees. The rectangular cell geometry (when α = 90 deg) could be one reason for this high value. However, this was seen only in FD1 and FD3, and the FD2 showed a significantly higher angle at this diameter. This has to be further verified with reliable computer simulations and more extensive test specimen sizes. The BF measurements prove to be reliable and repeatable for different stents. Both the RF_L_ and BF were reported earlier at specific diameters, and they can be further extended easily to measure at different diameters. We show that one can slightly modify a commonly available characterization setup like tensile/universal testing machine to obtain the data at varying diameters with good reliability.

To summarize, we applied and discussed different methods to characterize braided stents. This data can serve as guidelines for future design development when comparing to state-of-the-art. The bigger question unanswered is if the porosity, cell-area, and mechanical parameters play any role in the patient treatment outcomes. Generating such data sets and comparing it with long-term clinical performance could give us a better chance of understanding this question. To fulfill this, the data sets need to be as close to the real-life scenario as possible. In the current study, evaluating the porosity and cell-area in patient-derived models and measuring the mechanical forces in a range of diameters brings it a one-step closer to the real-case. Therefore, these methods serve as guidelines for further developments in the testing of braided FDs.

## 5. Limitations

Our study has a few shortcomings: 1) the porosity and cell-area were estimated based on one neuroradiologist’s deployment per device. The deployment itself and an aneurysm geometry can influence the discussed properties; thus, multiple deployments into the same geometry would be beneficial. However, the suggested method can be easily replicated with minimal effort for such a study. 2) More elaborate models that can accommodate cells growing on the deployed stent surface can give more insights about designs into inhibiting cell-growth along the aneurysms’ neck without using animal models.

Additionally, the other challenges are the reduced sample sizes, as only one FD per company was considered for the analysis. Therefore, the FDs were tested multiple times, and some defects in the material could have been introduced. Each device (even from the same manufacturer) can have slightly different mechanical properties, influencing the data. Thus, no conclusions about the variability between different stents of the same company could also be drawn. Ideally, the sample set of at least three devices has to be considered for each of the discussed experiments to avoid first, the damage of devices, and second, to ensure the reproducibility of the result over the different stents.

## 6. Conclusion

Laser microscopy of braided FD stents in 3D printed models of human vasculature allowed to access porosity and cell-area in realistic in-vitro conditions for the first time. 4D flow MRI allows, despite some artifacts, to access the flow diversion. The presented mechanical apparatus allows the consensus measurement of circumferential, longitudinal radial force, and bending force reliably and reproducibly as a function of diameter. We anticipate that these methods will add to the understanding of the stent properties and be adamant about developing new devices.

## Data Availability

The data that support the findings of this study are available from the lead author, PV, upon reasonable request.

## 7. Acknowledgments

This work was supported by the DFG from RTG “Materials4Brain” (GRK-2154); DFG under the cluster of excellence PMI 1267. Kiel University and the Medical Faculty are acknowledged for supporting the Molecular Imaging North Competence Center (MOIN CC) as a core facility for imaging in vivo. MOIN CC was founded by a grant from the European Regional Development Fund (ERDF) and the Zukunftsprogramm Wirtschaft of Schleswig-Holstein (Project no. 122-09-053). We are grateful to Phenox, Acandis, and Balt extrusion to provide the P64, DERIVO®, and Silk endovascular devices.

