## Supplementary material for "A realistic way to investigate the design, and mechanical properties of flow diverter stents": S. Figure 1; S. Table 1

1. **Frictional analysis of circumferential radial force device (RF_C_):**

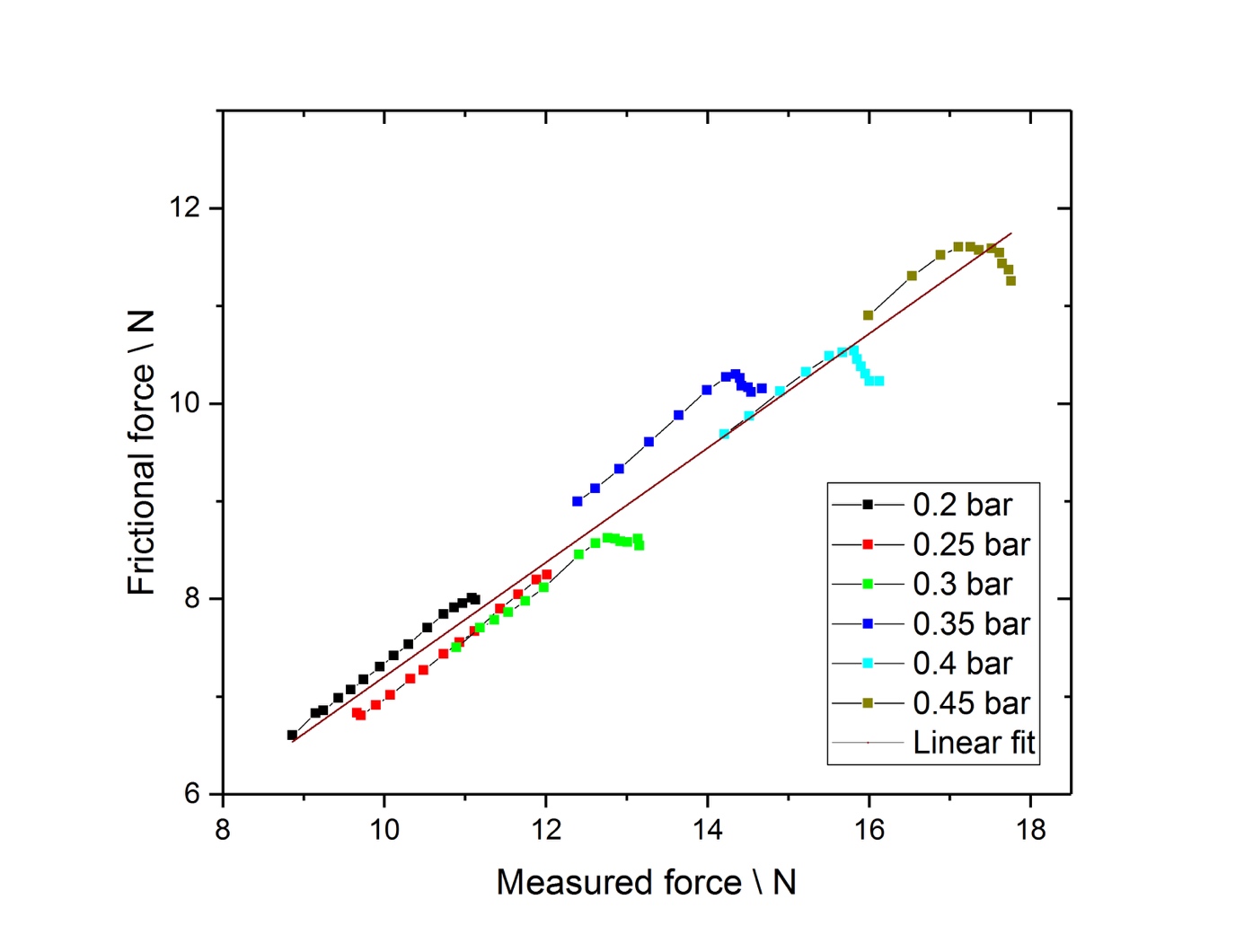
The dynamic friction between the radial blades plays an essential role in the measurements as it scales linearly with the measured radial force from the stents. When this is not accounted for, it results in a higher force than the device's actual force. Therefore, calibration was done by applying a known amount of force to the radial blades, and this was corrected with a correction factor (S. Figure 1). The linear fit in the figure has a slope of 0.59 with an intercept of 1.35.

S. Figure 1: Frictional analysis of the crimping analysis machine. As the stent expands in the machine, a force acts on the radial blades (Figure 2a), which creates friction between the radial plates during the analysis. If the friction is not taken into account, the analysis could be very misleading. To correct for friction, a known amount of force was applied (pressure) and was compared with the measured force. The image shows an approximate linear behavior, which one would expect for a dynamic friction scenario, the slope of the line being the coefficient of friction (0.59) with an intercept of 1.35 N for the considered apparatus. This also assumes that there is no friction between the stent and the radial blade itself.

2. **Flow analysis parameters:**

S. Table 1. Statistical parameters of velocity distribution cm/s in aneurysm sac and parental vessel with FD1-3 and without. The graphical representation of velocity distribution is shown in Figure 3 b-c. Abbreviations: Q1 – lower quantile, Q3 – upper quantile, SD – standard deviation, CV – coefficient of variance.

|  | Aneurysm sac | | | | Parental vessel | | | |
| --- | --- | --- | --- | --- | --- | --- | --- | --- |
| Absolute velocity | w/o FD | FD1 | FD2 | FD3 | w/o FD | FD1 | FD2 | FD3 |
| Min | 0.03 | 0.05 | 0.13 | 0.20 | 0.09 | 0.13 | 0.21 | 0.51 |
| Max | 78.29 | 101.83 | 136.31 | 82.87 | 112.79 | 121.14 | 128.24 | 109.08 |
| Mean | 20.53 | 8.76 | 10.35 | 8.71 | 34.89 | 36.03 | 41.27 | 36.12 |
| Q1 | 14.34 | 5.53 | 5.67 | 4.94 | 21.71 | 24.07 | 25.75 | 24.16 |
| Median | 20.74 | 6.89 | 7.42 | 6.45 | 35.21 | 36.67 | 42.14 | 35.55 |
| Q3 | 27.39 | 9.04 | 10.20 | 8.62 | 49.49 | 47.21 | 55.70 | 46.89 |
| SD | 13.53 | 8.45 | 11.32 | 9.02 | 20.12 | 17.54 | 21.33 | 16.74 |
| CV | 65.90 | 96.53 | 109.40 | 103.56 | 57.66 | 48.68 | 51.69 | 46.34 |

3. **Design parameters:**

S. Table 2: Calculated cell-area values for FD 1-3 at Pos. 1-3. The area was calculated by obtaining the dimensions of the cells using laser images (Figure 1e).

|  | Pos. 1 (*10^3^ µm^2^) | Pos. 2 (*10^3^ µm^2^) | Pos.3 (*10^3^ µm^2^) |
| --- | --- | --- | --- |
| FD 1 | 48.10 ± 11.01 | 41.66 ± 4.98 | 36.80 ± 3.69 |
| FD 2 | 78.02 ± 7.34 | 73.02 ± 7.02 | 68.62 ± 10.13 |
| FD 3 | 99.06 ± 11.79 | 92.30 ± 13.60 | 94.31 ± 8.68 |
